# Antibody response to SARS-CoV-2 mRNA vaccine in lung cancer patients: Reactivity to vaccine antigen and variants of concern

**DOI:** 10.1101/2022.01.03.22268599

**Authors:** Rajesh M Valanparambil, Jennifer Carlisle, Susanne L. Linderman, Akil Akthar, Ralph Linwood Millett, Lilin Lai, Andres Chang, Ashley A. McCook, Jeffrey Switchenko, Tahseen H. Nasti, Manpreet Saini, Andreas Wieland, Kelly E. Manning, Madison Ellis, Kathryn M. Moore, Stephanie L. Foster, Katharine Floyd, Meredith E. Davis-Gardner, Venkata-Viswanadh Edara, Mit Patel, Conor Steur, Ajay K. Nooka, Felicia Green, Margaret A. Johns, Fiona O’Brein, Uma Shanmugasundaram, Veronika I Zarnitsyna, Hasan Ahmed, Lindsay E. Nyhoff, Grace Mantus, Michael Garett, Srilatha Edupuganti, Madhusmita Behra, Rustom Antia, Jens Wrammert, Mehul S. Suthar, Madhav V. Dhodapkar, Suresh Ramalingam, Rafi Ahmed

**Affiliations:** Emory Vaccine Center, Emory University School of Medicine, Atlanta, GA; Department of Microbiology and Immunology, Emory University, Atlanta, GA; Winship Cancer Institute, Atlanta, GA; Department of Hematology and Medical Oncology, Emory University, Atlanta, GA; Department of Pediatrics, Emory University, Atlanta, GA; Yerkes National Primate Center, Atlanta, GA; ICGEB-Emory Vaccine Centre, International Centre for Genetic Engineering and Biotechnology (ICGEB), Aruna Asaf Ali Marg, New Delhi, India; Department of Biology, Emory University, Atlanta, GA; Center for Childhood Infections and Vaccines of Children’s Healthcare of Atlanta, Department of Pediatrics, Emory University School of Medicine, Atlanta, GA; Hope Clinic of Emory Vaccine Center, Emory University School of Medicine, Atlanta, GA; Department of Otolaryngology, The Ohio State University, Columbus, Ohio; Pelotonia Institute for Immuno-Oncology, The Ohio State University, Columbus, Ohio; Department of Biostatistics and Bioinformatics, Rollins School of Public Health, Emory University; Biostatistics Shared Resource, Winship Cancer Institute, Emory University

## Abstract

**Purpose:** We investigated SARS-CoV-2 mRNA vaccine-induced binding and live-virus neutralizing antibody response in NSCLC patients to the SARS-CoV-2 wild type strain and the emerging Delta and Omicron variants.

**Methods:** 82 NSCLC patients and 53 healthy adult volunteers who received SARS-CoV-2 mRNA vaccines were included in the study. Blood was collected longitudinally, and SARS-CoV-2-specific binding and live-virus neutralization response to 614D (WT), B.1.617.2 (Delta), B.1.351 (Beta) and B.1.1.529 (Omicron) variants were evaluated by Meso Scale Discovery (MSD) assay and Focus Reduction Neutralization Assay (FRNT) respectively. We determined the longevity and persistence of vaccine-induced antibody response in NSCLC patients. The effect of vaccine-type, age, gender, race and cancer therapy on the antibody response was evaluated.

**Results:** Binding antibody titer to the mRNA vaccines were lower in the NSCLC patients compared to the healthy volunteers (P=<0.0001). More importantly, NSCLC patients had reduced live-virus neutralizing activity compared to the healthy vaccinees (P=<0.0001). Spike and RBD-specific binding IgG titers peaked after a week following the second vaccine dose and declined after six months (P=<0.001). While patients >70 years had lower IgG titers (P=<0.01), patients receiving either PD-1 monotherapy, chemotherapy or a combination of both did not have a significant impact on the antibody response. Binding antibody titers to the Delta and Beta variants were lower compared to the WT strain (P=<0.0001). Importantly, we observed significantly lower FRNT_50_ titers to Delta (6-fold), and Omicron (79-fold) variants (P=<0.0001) in NSCLC patients.

**Conclusions:** Binding and live-virus neutralizing antibody titers to SARS-CoV-2 mRNA vaccines in NSCLC patients were lower than the healthy vaccinees, with significantly lower live-virus neutralization of B.1.617.2 (Delta), and more importantly, the B.1.1.529 (Omicron) variant compared to the wild-type strain. These data highlight the concern for cancer patients given the rapid spread of SARS-CoV-2 Omicron variant.

## INTRODUCTION

Coronaviruses are a group of enveloped, single stranded RNA viruses that infect vertebrates. Human coronavirus (HCoV) infection causes mild to severe respiratory disease in humans. Prior to December 2019, of the six known HCoVs infections, severe acute respiratory syndrome coronavirus (SARS-CoV) and Middle East respiratory syndrome coronavirus (MERS-CoV) infections resulted in severe respiratory disease outbreaks with very high mortality rates^1,2^. In December 2019, an increase in the cases of severe respiratory illness was reported from Wuhan, China. SARS-CoV-2 was identified as the virus causing the disease and the illness was termed as COVID-19. By March 2021 the disease had spread all over the world making it a global pandemic^3,4^. As of November 2021, more than 5 million people have succumbed to SARS-CoV-2 with the highest mortality rate among elderly people. In December 2020 two vaccines, BNT162b2 by Pfizer and mRNA-1273 by Moderna were made available to the general public in the U.S. Both the vaccines had more than 95% efficacy in controlling SARS-CoV-2 infection^5,6^. However, since then several variants of concerns (VOCs) have emerged resulting in breakthrough infections. Notably the B.1.617.2 (Delta) variant accounts for more than 90% of the COVID-19 cases in the U.S. Nevertheless, both the vaccines have proved to reduce hospitalization and death remarkably compared to the unvaccinated individuals^7,8^.

Approximately 2 million patients are diagnosed with lung cancer every year globally; it is the leading cause of cancer-related deaths with nearly 1.76 million deaths per year. The median age of lung cancer diagnosis is 70 years. Non-small cell lung carcinoma (NSCLC) accounts for 84% of all the lung cancer diagnoses^9,10^. Immune dysregulation is a common phenomenon in cancer patients due to tumor malignancy and immunomodulatory therapies that the patients receive. It is thus important to evaluate the efficacy of SARS-CoV-2 vaccination in these patients ^11 12,13^. A recent study in thoracic cancer patients receiving the BNT162b2 vaccine (99.3%) has shown to be efficient in generating protective antibody response ^14^. However, this study did not show the efficacy of vaccinated NSCLC patients to neutralize emerging variants of concern (VOC). Since late 2020, several SARS-CoV-2 variants of concern have emerged. These variants have acquired mutations that impact virus transmission and pathogenicity. Some of these variants are poorly neutralized by postvaccination serum from healthy individuals ^15^. In November 2021, the SARS-CoV-2 Omicron variant which carries 32 mutations in its spike protein was identified in South Africa and has since then spread to several countries in the world and have become a dominant SARS-CoV-2 strain worldwide ^16,17^ The Omicron variant escapes both vaccine-induced and therapeutic antibodies in healthy individuals but can be neutralized by booster dose-induced antibody response ^17^. Here we studied the efficacy of mRNA vaccination in NSCLC patients to generate neutralizing antibodies against the rapidly evolving SARS CoV-2 B.1.1.529 Omicron variant and the previously dominant B.1.617.2 (Delta) variants.

## METHODS

### NSCLC patient cohort

Peripheral blood samples from NSCLC patients were collected at Winship Cancer Institute following written informed consent approved by the Institutional Review Board at Emory University. Blood samples were processed to isolate plasma and mononuclear cells. The patient demographics for all 82 NSCLC patients enrolled in the study are in Table 1. Clinical characteristics and treatment information are tabulated in Table 1a, and the COVID mRNA vaccine type is described in Table 1b.

**Table 1 -.**
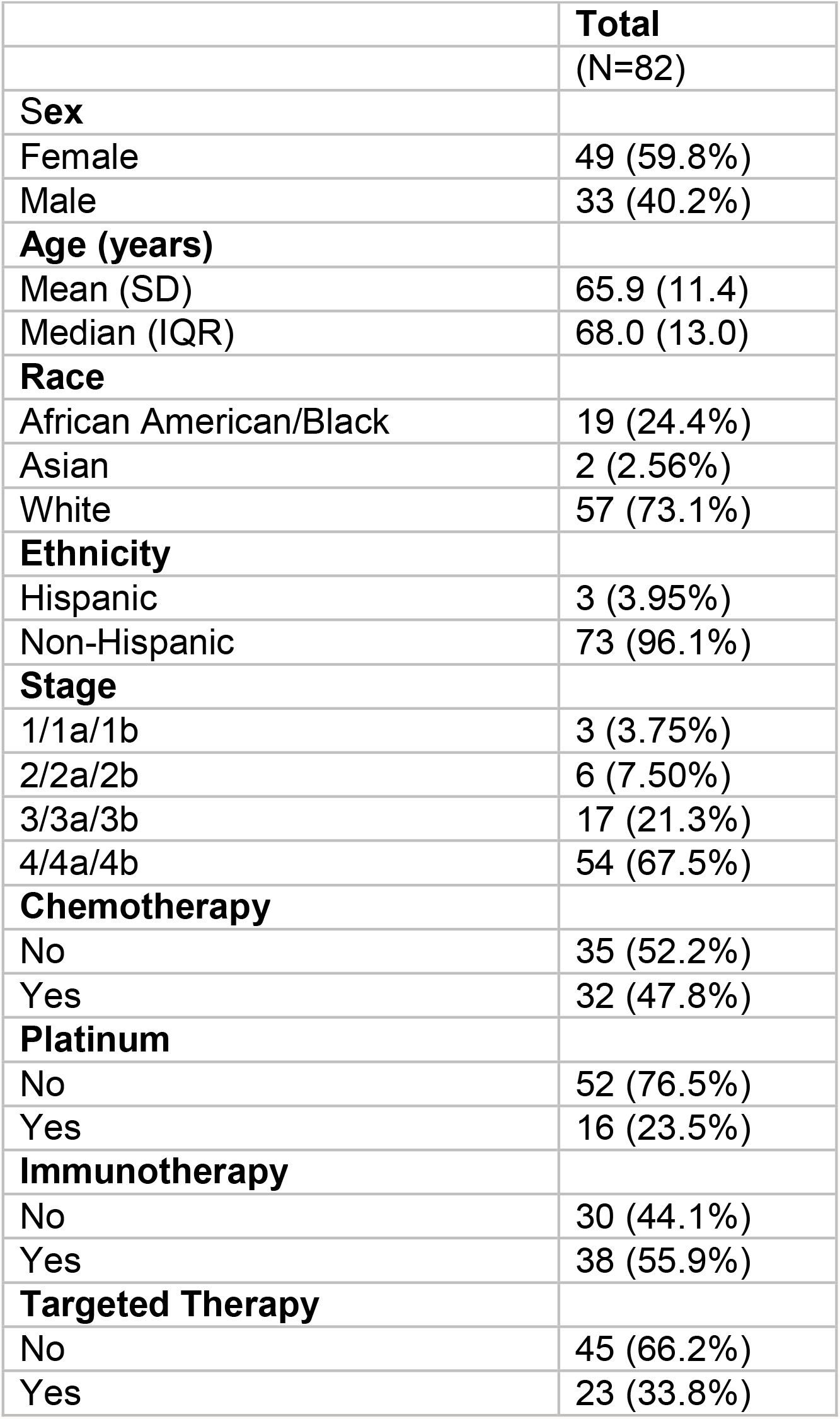
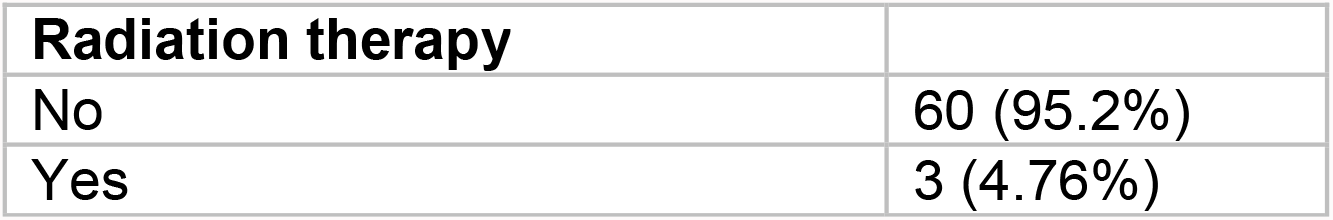
Descriptive statistics of patient characteristics.

|  | **Total** |
| --- | --- |
|  | (N=82) |
| **Sex** |  |
| Female | 49 (59.8%) |
| Male | 33 (40.2%) |
| **Age (years)** |  |
| Mean (SD) | 65.9 (11.4) |
| Median (IQR) | 68.0 (13.0) |
| **Race** |  |
| African American/Black | 19 (24.4%) |
| Asian | 2 (2.56%) |
| White | 57 (73.1%) |
| **Ethnicity** |  |
| Hispanic | 3 (3.95%) |
| Non-Hispanic | 73 (96.1%) |
| **Stage** |  |
| 1/1a/1b | 3 (3.75%) |
| 2/2a/2b | 6 (7.50%) |
| 3/3a/3b | 17 (21.3%) |
| 4/4a/4b | 54 (67.5%) |
| **Chemotherapy** |  |
| No | 35 (52.2%) |
| Yes | 32 (47.8%) |
| **Platinum** |  |
| No | 52 (76.5%) |
| Yes | 16 (23.5%) |
| **Immunotherapy** |  |
| No | 30 (44.1%) |
| Yes | 38 (55.9%) |
| **Targeted Therapy** |  |
| No | 45 (66.2%) |
| Yes | 23 (33.8%) |
| **Radiation therapy** |  |
| No | 60 (95.2%) |
| Yes | 3 (4.76%) |

**Table 2 -.**
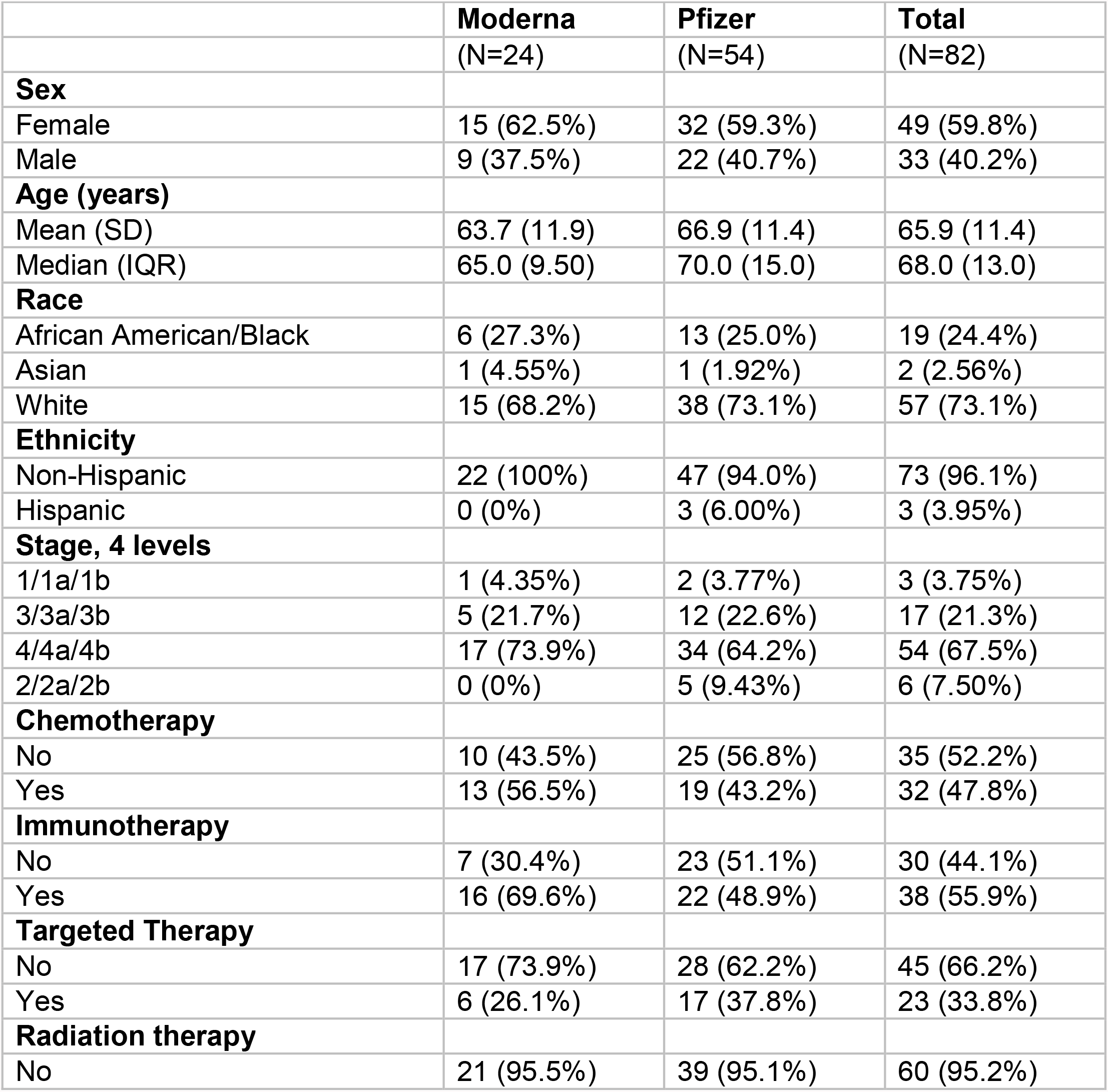
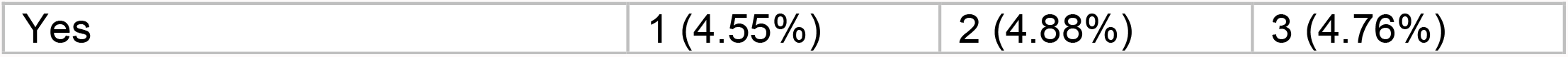
Descriptive statistics by patient’s COVID vaccine type.

|  | **Moderna** | **Pfizer** | **Total** |
| --- | --- | --- | --- |
|  | (N=24) | (N=54) | (N=82) |
| **Sex** |  |  |  |
| Female | 15 (62.5%) | 32 (59.3%) | 49 (59.8%) |
| Male | 9 (37.5%) | 22 (40.7%) | 33 (40.2%) |
| **Age (years)** |  |  |  |
| Mean (SD) | 63.7 (11.9) | 66.9 (11.4) | 65.9 (11.4) |
| Median (IQR) | 65.0 (9.50) | 70.0 (15.0) | 68.0 (13.0) |
| **Race** |  |  |  |
| African American/Black | 6 (27.3%) | 13 (25.0%) | 19 (24.4%) |
| Asian | 1 (4.55%) | 1 (1.92%) | 2 (2.56%) |
| White | 15 (68.2%) | 38 (73.1%) | 57 (73.1%) |
| **Ethnicity** |  |  |  |
| Non-Hispanic | 22 (100%) | 47 (94.0%) | 73 (96.1%) |
| Hispanic | 0 (0%) | 3 (6.00%) | 3 (3.95%) |
| **Stage, 4 levels** |  |  |  |
| 1/1a/1b | 1 (4.35%) | 2 (3.77%) | 3 (3.75%) |
| 3/3a/3b | 5 (21.7%) | 12 (22.6%) | 17 (21.3%) |
| 4/4a/4b | 17 (73.9%) | 34 (64.2%) | 54 (67.5%) |
| 2/2a/2b | 0 (0%) | 5 (9.43%) | 6 (7.50%) |
| **Chemotherapy** |  |  |  |
| No | 10 (43.5%) | 25 (56.8%) | 35 (52.2%) |
| Yes | 13 (56.5%) | 19 (43.2%) | 32 (47.8%) |
| **Immunotherapy** |  |  |  |
| No | 7 (30.4%) | 23 (51.1%) | 30 (44.1%) |
| Yes | 16 (69.6%) | 22 (48.9%) | 38 (55.9%) |
| **Targeted Therapy** |  |  |  |
| No | 17 (73.9%) | 28 (62.2%) | 45 (66.2%) |
| Yes | 6 (26.1%) | 17 (37.8%) | 23 (33.8%) |
| **Radiation therapy** |  |  |  |
| No | 21 (95.5%) | 39 (95.1%) | 60 (95.2%) |
| Yes | 1 (4.55%) | 2 (4.88%) | 3 (4.76%) |

### Healthy control cohort

A total of 53 healthy volunteers vaccinated with COVID mRNA vaccines were recruited, and samples were collected at the Hope clinic, Atlanta following written informed consent approved by the Institutional Review Board at Emory University. Of these, 22 volunteers received mRNA-1273 and 31 received BNT162b2.

### MesoScale Discovery Assay

V-PLEX COVID-19 Respiratory Panel 2 Kit (K15372U panel 2) were used to measure the IgG, IgM and IgA antibody against the antigens SARS-CoV-2 Spike, receptod binding domain (RBD), N-terminal domain (NTD) and nucleocapsid, following the manufacturer’s recommendations. To measure spike-specific IgG against SARS-CoV-2 variants, V-PLEX COVID-19 Respiratory Panel 13 and panel 14 Kits (K15463U and K15468U) were used. To assess IgG binding, plasma samples were diluted 1:5000 or 1:25,000 and MSD Reference Standard-1 was diluted per MSD instructions in MSD Diluent 100. 50 uL of each sample and Reference Standard-1 dilution was added to the plates and incubated for two hours at room temperature, shaking at a speed of 700 rpm. Following this, 50 uL per well of 1X MSD SULFO-TAG™ Anti-Human IgG Antibody was added and incubated for one hour at room temperature, shaking at a speed of 700 rpm. Following the detection reagent step, 150 uL per well of MSD Gold™ Read Buffer B was added to each plate immediately prior to reading on an MSD plate reader. Plates were washed three times with 150 uL of wash solution B provided in the kit between each step. The limit of detection was defined as 1,000 RLU for each assay.

### Viruses and cells

VeroE6 cells were obtained from ATCC (clone E6, ATCC, #CRL-1586) and cultured in complete DMEM medium consisting of 1x DMEM (VWR, #45000-304), 10% FBS, 25mM HEPES Buffer (Corning Cellgro), 2mM L-glutamine, 1mM sodium pyruvate, 1x Non-essential Amino Acids, and 1x antibiotics. VeroE6-TMPRSS2 cells were generated and cultured as previously described^18^. nCoV/USA_WA1/2020 (WA/1), closely resembling the original Wuhan strain and resembles the spike used in the mRNA-1273 and Pfizer-BioNTech vaccine, was propagated from an infectious SARS-CoV-2 clone as previously described^19^. icSARS-CoV-2 was passaged once to generate a working stock. The B.1.351 variant isolate (hCoV-19/USA/MD-HP01542/2021), kindly provided by Dr. Andy Pekosz (John Hopkins University, Baltimore, MD), was propagated once in VeroE6-TMPRSS2 cells to generate a working stock. hCoV19/EHC_C19_2811C (referred to as the B.1.1.529 variant) was derived from a mid-turbinate nasal swab collected in December 2021^20^. This SARS-CoV-2 genome is available under GISAID accession number EPI_ISL_7171744. Using VeroE6-TMPRSS cells, the B.1.1.529 variant was plaque purified directly from the nasal swab, propagated once in a 12-well plate, and expanded in a confluent T175 flask to generate a working stock. All viruses used in this study were deep sequenced and confirmed as previously described^18^.

### Focus Reduction Neutralization Assay

FRNT-mNG assays were performed on VeroE6 cells and FRNT assays were performed on Vero-TMPRSS2 cells as previously described^18,21,22^. Samples were diluted at 3-fold in 8 serial dilutions using DMEM (VWR, #45000-304) in duplicates with an initial dilution of 1:10 in a total volume of 60 μl. Serially diluted samples were incubated with an equal volume of icSARS-CoV-2-mNG, WA1/2020, B.1.351, or B.1.1.529 (100-200 foci per well based on the target cell) at 37° C for 45 minutes in a round-bottomed 96-well culture plate. The antibody-virus mixture was then added to cells and incubated at 37°C for 1 hour. Post-incubation, the antibody-virus mixture was removed and 100 µl of pre-warmed 0.85% methylcellulose (Sigma-Aldrich, #M0512-250G) overlay was added to each well. Plates were incubated at 37° C for 18 hours and the methylcellulose overlay was removed and washed six times with PBS. For Omicron infections, cells were incubated for 40-48h. Cells were fixed with 2% paraformaldehyde in PBS for 30 minutes. Following fixation, plates were washed twice with PBS and permeabilization buffer (0.1% BSA [VWR, #0332], Saponin [Sigma, 47036-250G-F] in PBS) was added to permeabilized cells for at least 20 minutes. Cells were incubated with either an anti-SARS-CoV spike primary antibody directly conjugated to Alexaflour-647 (CR3022-AF647) or an anti-SARS-CoV spike primary antibody directly conjugated to biotin (CR3022-biotin) for at least 4 hours at room temperature. For CR3022-AF647, cells were washed three times in PBS and foci were visualized on an ELISPOT reader (CTL). For CR3022-biotin, cells were washed three times in PBS and avidin-HRP was added for 1 hour at room temperature followed by three washes in PBS. Foci were visualized using TrueBlue HRP substrate (KPL, # 5510-0050) and imaged on an ELISPOT reader (CTL). Antibody neutralization was quantified by counting the number of foci for each sample using the Virodot program^23^. The neutralization titers were calculated as follows: 1 - (ratio of the mean number of foci in the presence of sera and foci at the highest dilution of respective sera sample). Each specimen was tested in duplicates. Samples that did not neutralize at the limit of detection at 50% are plotted at 12 and was used for geometric mean calculations.

### Statistical analysis

Statistical analysis was conducted using Graphpad Prism V9 and R 4.1.2. For all analyses, significance level was set at P <0.05, two-tailed. Descriptive statistics were performed to tabulate patients’ demographic and clinical characteristics by COVID vaccine type. Frequency and percentage, mean and standard deviation or median with interquartile range were reported based on the data structure of variable (categorical vs continuous). Statistical differences were assessed by group using one-way analysis of variance, Kruskal-Wallis test, student’s t-test, or Mann-Whitney test. The strength of association between laboratory biomarkers were tested with Pearson or Spearman test. Further univariate analysis was conducted with simple linear regression on patients’ assays collected in the study. Appropriate statistical tests were selected by validity of assumptions the variables in analyses. The plots of the residuals (Q-Q plots) from each variable were used to examine whether parametric or non-parametric statistical test would be utilized. Multiple comparisons are accounted for in our statistical tests comparing different groups in the study.

## RESULTS

### Binding antibody response to SARS-CoV-2 mRNA vaccines in NSCLC patients

First, we evaluated the binding antibody response to the mRNA vaccines in NSCLC patients. Plasma collected from healthy mRNA vaccine recipients were used as controls. Pre-pandemic plasma samples collected from healthy individuals were used to set the detection limit of the assay. A month after the second dose of vaccination, most NSCLC patients showed a strong binding IgG, IgA and IgM response to the mRNA vaccines. However, spike, RBD and NTD-specific IgG titers were significantly lower (P=<0.0001 for spike, 0.0002 for RBD and <0.0001 for NTD) compared to the healthy controls. As SARS-CoV-2 nucleocapsid is not a component of the mRNA vaccine, patients with a history of SARS-CoV-2 infection were identified by the presence of high anti-nucleocapsid titer (N+) in their plasma. These N+ patients had higher levels of spike, RBD and NTD-specific antibody compared to the SARS-CoV-2 naïve NSCLC patients (P=<0.0001) (Fig 1A-C). Next, we examined the presence of vaccine specific IgA titers in the plasma of NSCLC patients. Similar to the IgG titers, vaccine-specific IgA titers were lower (P=0.0001 for spike, <0.0001 for RBD and <0.0001 for NTD) in the NSCLC patients compared to the healthy controls. Patients who were N+ had higher IgA titers compared to SARS-CoV-2 naïve NSCLC patients (P=0.0136 for spike, 0.0008 for RBD and 0.0027 for NTD) (Fig 1D-E). Like the IgG and IgA titers, we observed lower vaccine-specific IgM titers in the plasma of NSCLC patients compared to the healthy individuals (P=<0.0001 for spike, 0.0002 for RBD and 0.0005 for NTD). We did not notice a significant difference in the spike and RBD-specific IgM titer in N+ NSCLC patients compared to the SARS-CoV-2 naïve patients. NTD-specific IgM was higher (P=0.0349) in the N+ patients to the SARS-CoV-2 naïve NSCLC patients (Fig 1G-I). Next, we examined if the Spike-specific antibody titers correlate with the RBD and NTD-specific antibody levels. As shown in supplementary. figure 1 A-C (online only), the binding spike-specific IgG, IgA and IgM titers strongly correlate with the RBD-specific titers (R^2^=0.9616 for IgG, R^2^=0.7697 for IgA and R^2^=0.6495 for IgM). Similarly, the spike-specific antibody titers also correlate with NTD-specific titers (R^2^=0.9659 for IgG, R^2^=0.7797 for IgA and R^2^=0.7644 for IgM) (supplementary figure 1 D-F. online only). Taken together, the data show that, mRNA vaccination induced vaccine-specific binding antibody titers in NSCLC patients, however the antibody titer was lower than the titers in healthy individuals. Spike-specific antibody titers strongly correlated with RBD and NTD titers.

**Figure 1.**
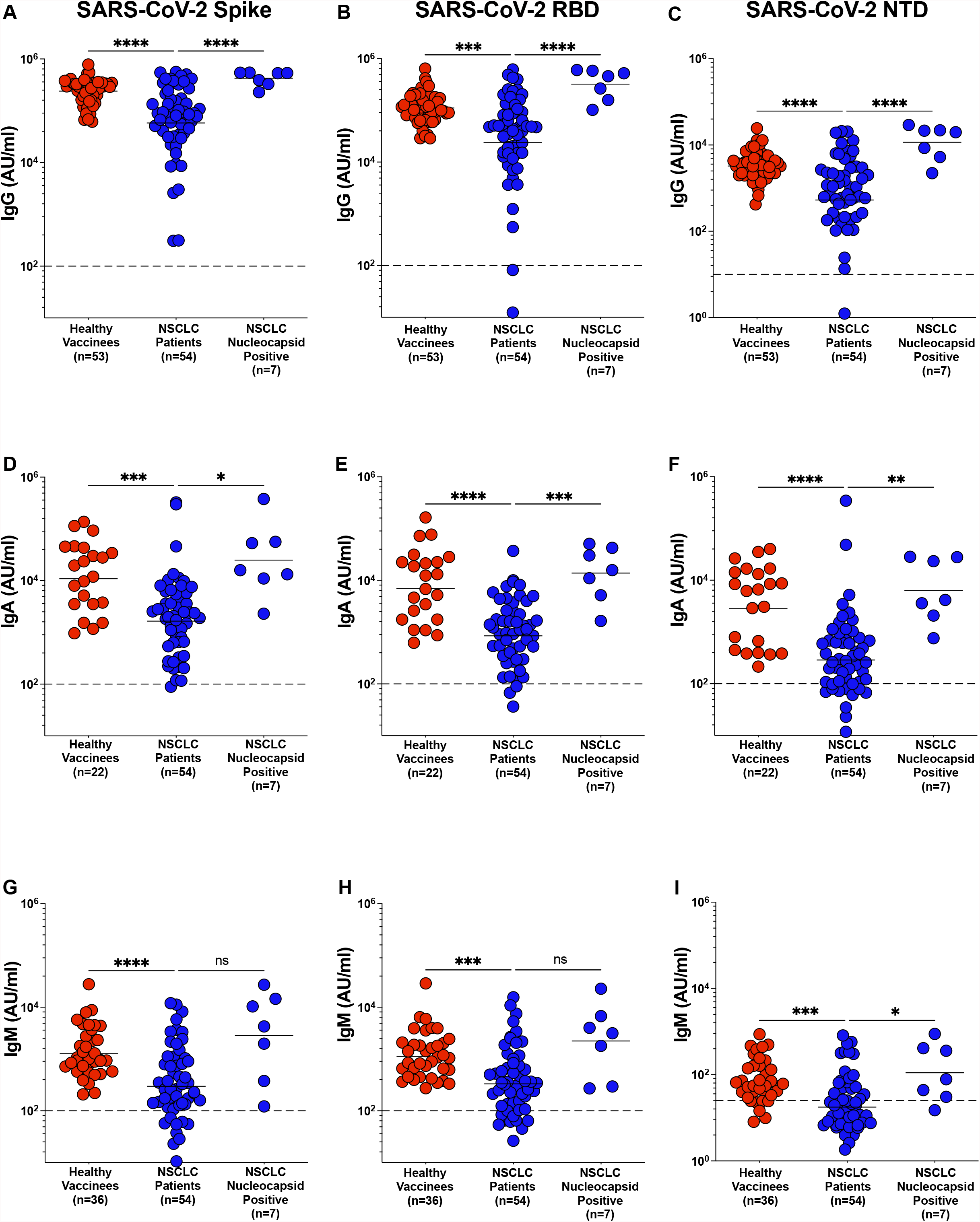
Antibody response to SARS-CoV-2 mRNA vaccine in NSCLC patients. Figure 1 A – I. Spike, RBD and NTD specific IgG (Figure 1A-C), IgA (Figure 1D-F) and IgM (Figure 1G-I), titers in plasma from healthy vaccinees, NSCLC patients and NSCLC patients with prior exposure to SARS-CoV-2 infection was measured within two months after the second dose of mRNA vaccination. Pre-pandemic plasma samples from healthy individuals was used to set the detection limit for IgG, IgA and IgM titers. Statistical differences were measured using one-way anova. Graph shows the mean and s.e.m. ns not significant, *p≤0.05, **p≤0.01, ***p≤0.001, ****p≤0.0001.

**Figure 2.**
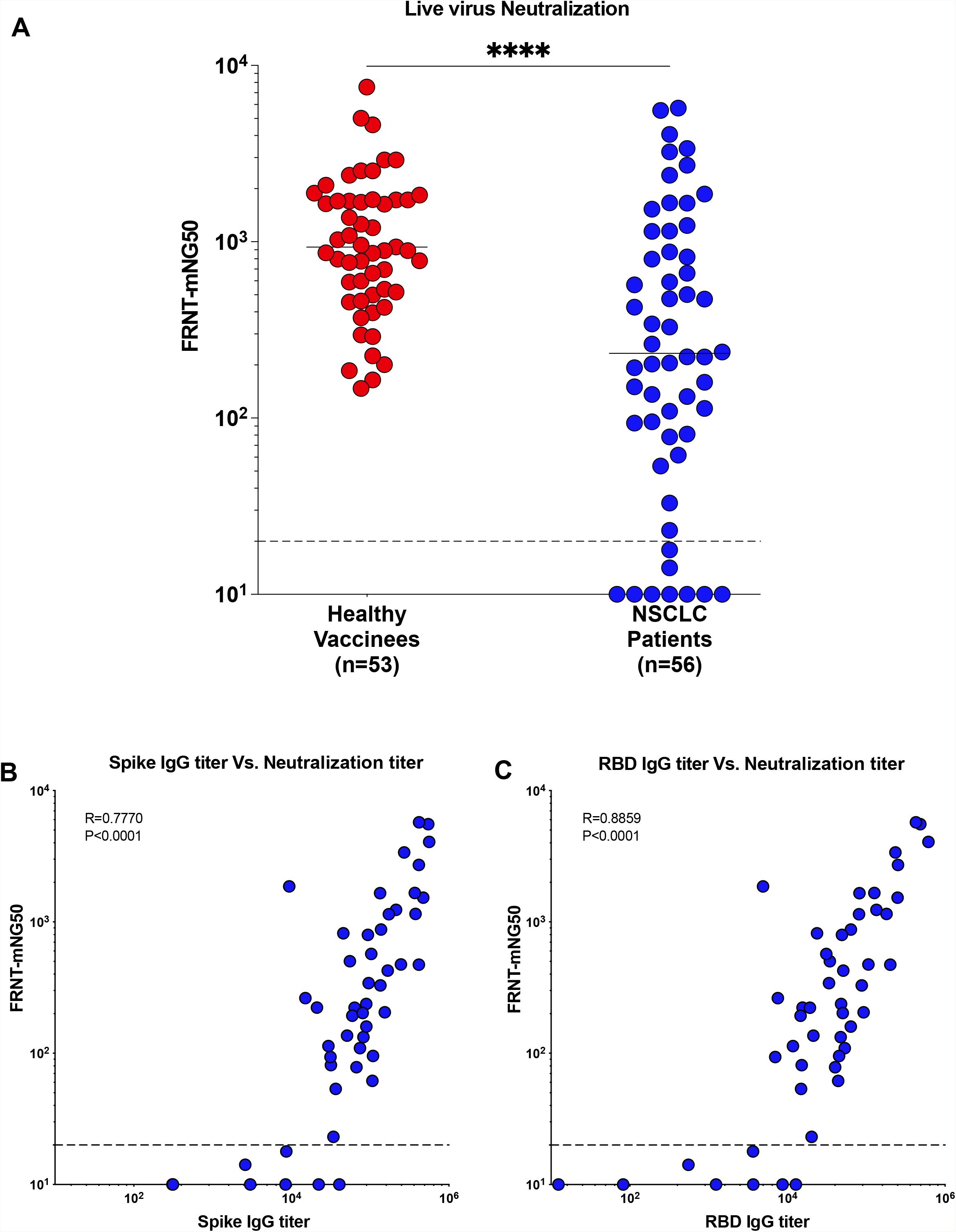
Live virus neutralizing antibody response correlates with the binding response. Figure 2A. Live virus neutralization of WT (614D) virus by sera from NSCLC patients compared to the samples from healthy vaccinees. Figure 2 B. Correlation of spike-specific IgG titer and FRNT_50_ titer. Figure 2 Correlation between RBD-specific IgG titer and FRNT_50_ titer. Simple linear regression analysis was performed to do correlation analysis and p-values were obtained from Pearson r correlation method. Statistical differences were measured using one-way anova. Graph shows the mean and s.e.m. ****p≤0.0001.

### Neutralizing antibody response to SARS-CoV-2 mRNA vaccination in NSCLC patients

Next, we performed a live virus neutralization assay to determine the presence of neutralizing antibodies against the WA1\2020 stain in healthy vaccinees and NSCLC patients in response to mRNA vaccination. Like the binding antibody titers, the neutralizing antibody titers in the plasma of NSCLC patients were significantly lower compared to the healthy vaccinees (P=<0.0001). Though most of the NSCLC patients generated neutralizing antibodies, a subset of these patients failed to neutralize live virus (Fig 3A). The focus reduction neutralization test (FRNT)_50_ for live virus correlated with the binding spike antibody titer (P=<0.0001) (R=0.7770 for Spike-IgG) (Fig 3B). Similarly, we also observed the FRNT_50_ also correlated with RBD specific IgG titers in NSCLC patients ((P=<0.0001) (R=0.8859 for RBD-IgG). Taken together, these data demonstrate that in response to vaccination most NSCLC patients generate detectable neutralizing antibody titers, albeit at lower levels compared to the healthy vaccinees. However, a significant fraction of NSCLC patients (9 out of 54) failed to mount a detectable neutralizing antibody response

**Figure 3.**
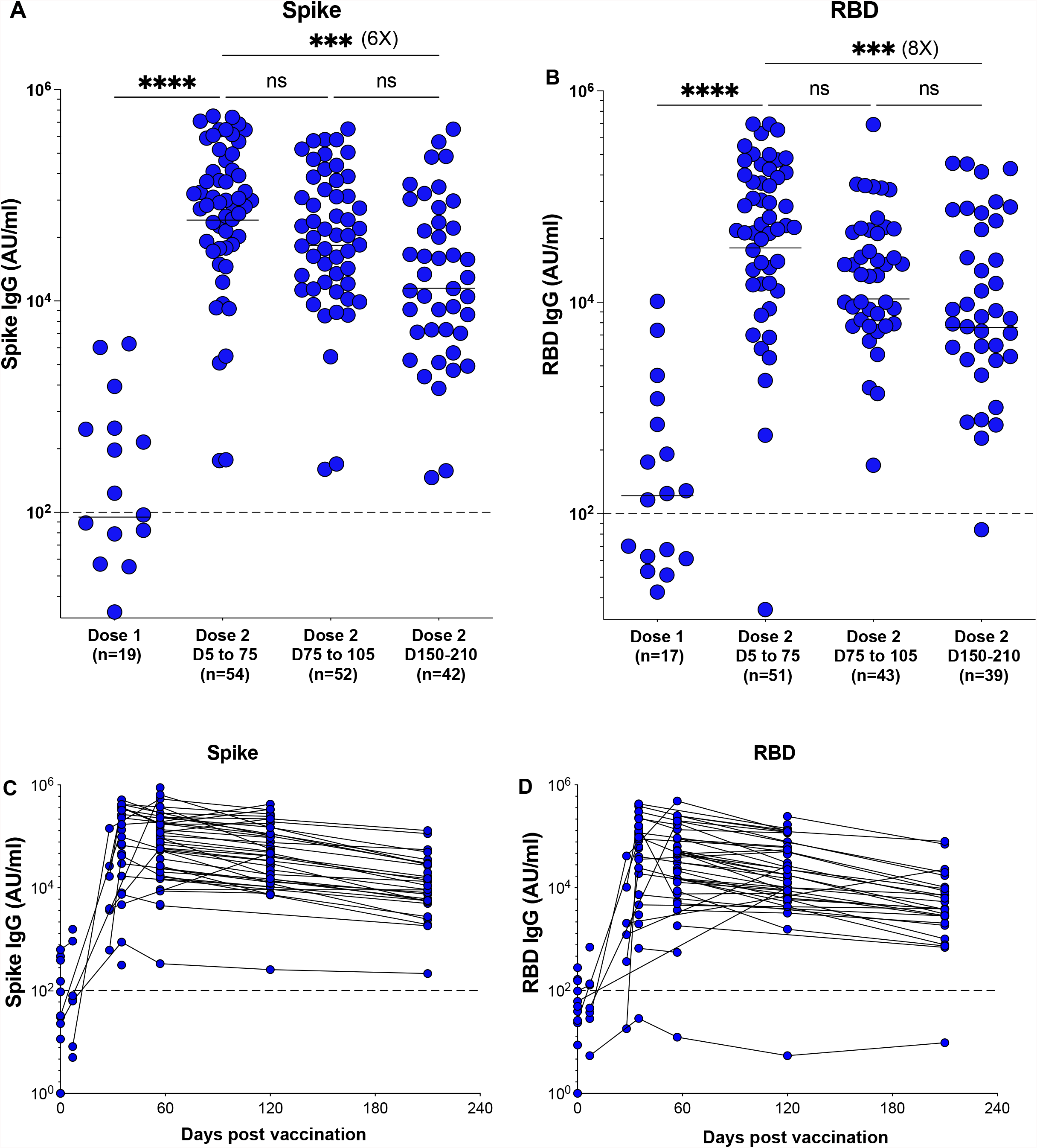
Longevity of anti-spike and anti-RBD antibody titers. Spike (Figure 3A and C) and RBD (Figure 3B and D) specific IgG titers in NSCLC patients measured at week 1-3 after the first dose, at 1-2 weeks, 1-2 months, 3-4 months and 5-6 months after the second dose of vaccination in NSCLC patients. Pre-pandemic plasma samples used in figure 1 was used to set the detection limit titer. Statistical differences were measured using one-way anova. Graph shows the mean and s.e.m. ns not significant, *p≤0.05, ***p≤0.001, ****p≤0.0001.

### Longitudinal analysis of spike and RBD-specific antibody response in vaccinated NSCLC patients

To evaluate the persistence of the vaccine-specific antibody response, we longitudinally measured the binding antibody response to mRNA vaccines in our NSCLC patients over six months. Anti-Spike and anti-RBD IgG titers peaked after a week following the second vaccine dose (P=<0.0001). Both anti-spike and anti-RBD IgG titers tended to decrease approximately 3 months after the second dose, although this did not reach statistical significance. However, there was a significant decrease in both anti-spike (6-fold) (P=<0.0003) and anti-RBD specific (8-fold) (P=<0.0009) binding antibody responses at 6 months following the second dose of vaccination compared to the respective peak IgG titers (Fig 3A to D).

### Impact of patient baseline characteristics on vaccine response

We examined if the demographic characteristics of our NSCLC cohort influenced the antibody response to vaccination. We observed a significant decrease in the binding spike-specific IgG titer in elderly (>70 years old) NSCLC patients compared to the patients who were <60 years of age (P=<0.0034) (Fig 4A). We did not see a significant difference in the binding IgG titers between the male and female patients (Fig 4B). However, we noticed an increase in spike specific IgG titer in the African American population compared to the white patients (p=0.0235) (Fig 4C). As many of the NSCLC patients in our cohort are actively receiving cancer therapy, we evaluated if the difference in the kind cancer therapy had an influence on the antibody response to vaccination. Based on the type of therapy, the NSCLC cohort was divided into five subsets (patients receiving PD-1 targeted therapy (n=15), PD-1 and chemotherapy (n=10) chemotherapy (n=6) and other targeted therapy (n=6)). At the time of SARS-CoV-2 vaccination, 13 patients were receiving no therapy. We did not see a notable difference in the binding spike-specific IgG titers among patients receiving different cancer therapies compared to patients receiving no therapy (Fig 4D). Taken together, these data suggest that while age and race of the patient influenced the antibody response to vaccination, gender and cancer therapy did not have any effect on the antibody response to SARS-CoV2 vaccination in our NSCLC cohort.

**Figure 4.**
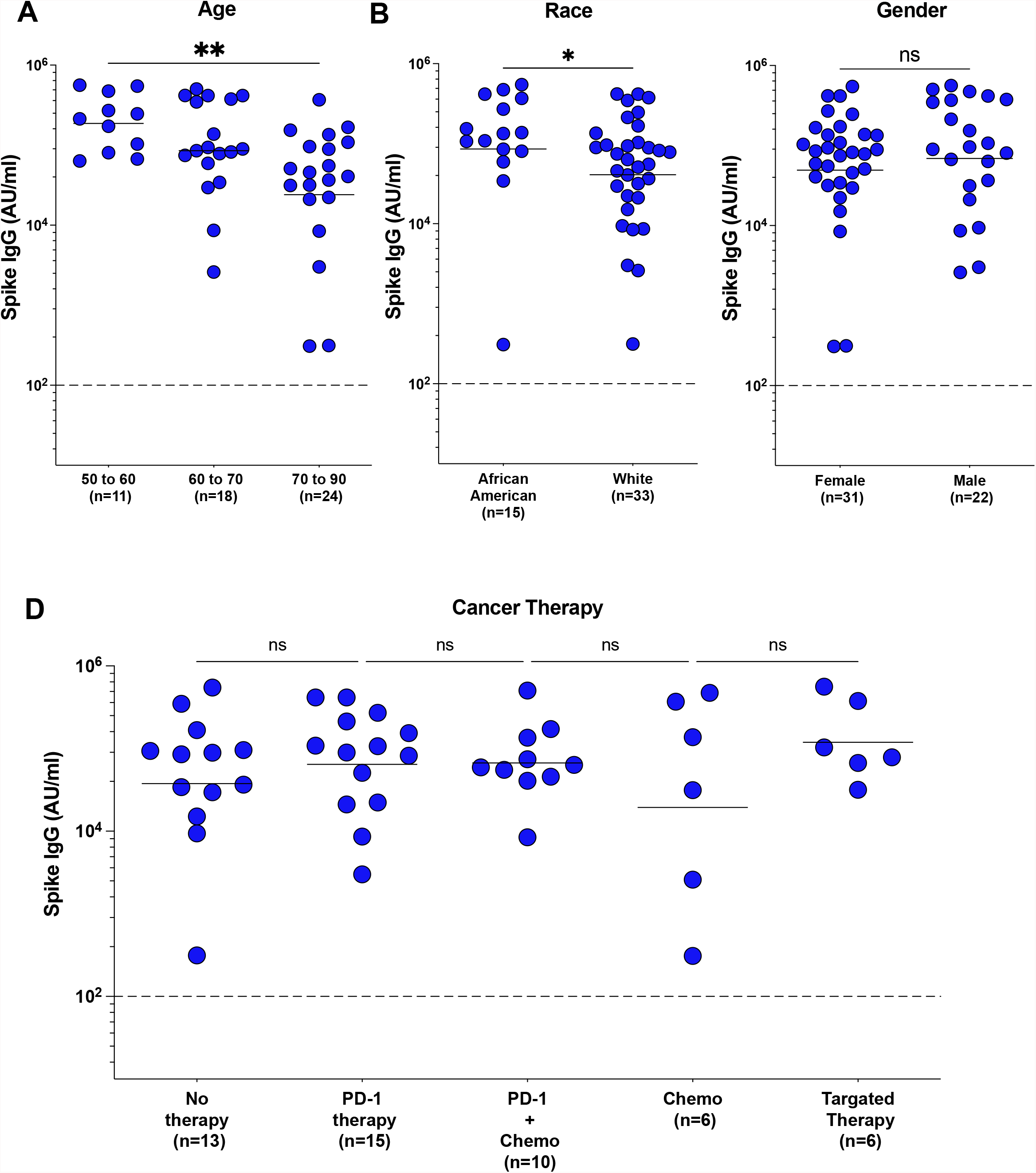
Influence of demographic factors on the antibody response to mRNA vaccination in NSCLC patients. Figure 4A. NSCLC patients were divided into three age groups (age 50-60, 60-70 and 70-90) and their binding anti-spike IgG titers were determined. Figure 4B. Spike specific IgG titers in African American and White NSCLC patients. Figure 4C. Binding anti-spike IgG titers in male and female NSCLC patients. Figure 4D. Effect of different cancer therapies (no therapy, PD-1 therapy, PD-1 with chemotherapy, chemotherapy, and targeted therapy) on the spike-specific IgG titers in response to the mRNA vaccines. Statistical differences were measured using Mann-Whitney test and one-way anova where applicable. Graph shows the mean and s.e.m. *p≤0.05, **p≤0.01.

### Antibody titer in NSCLC patients against SARS-CoV-2 variants

Next, we analyzed the spike-specific IgG titers in the plasma of NSCLC patients against different variants. Fig 5A, shows the median spike-specific IgG titers in the plasma of both healthy vaccinees (n=52) and the NSCLC patients (n=54) to 15 different SARS-CoV-2 variants. We observed highly variable IgG titers among the different variants. While the spike-specific IgG titer was the highest for the WT (614D) spike, the titer to the spike protein of the SARS-CoV-2 B.1.351 (Beta) variant was the lowest (Fig 5A). Spike-specific IgG titer to the B.1.617.2 (Delta) and the Beta variant were significantly lower compared to the titer against the WT spike (P=<0.0001) in healthy vaccinees (Fig 5B). Similar to the observations in the healthy vaccinees, spike-specific IgG titer to the Delta variant (P=<0.0001) and the Beta variant (P=0.0026) was lower compared to the titer against the WT spike in NSCLC patients (Fig 5C).

**Figure 5.**
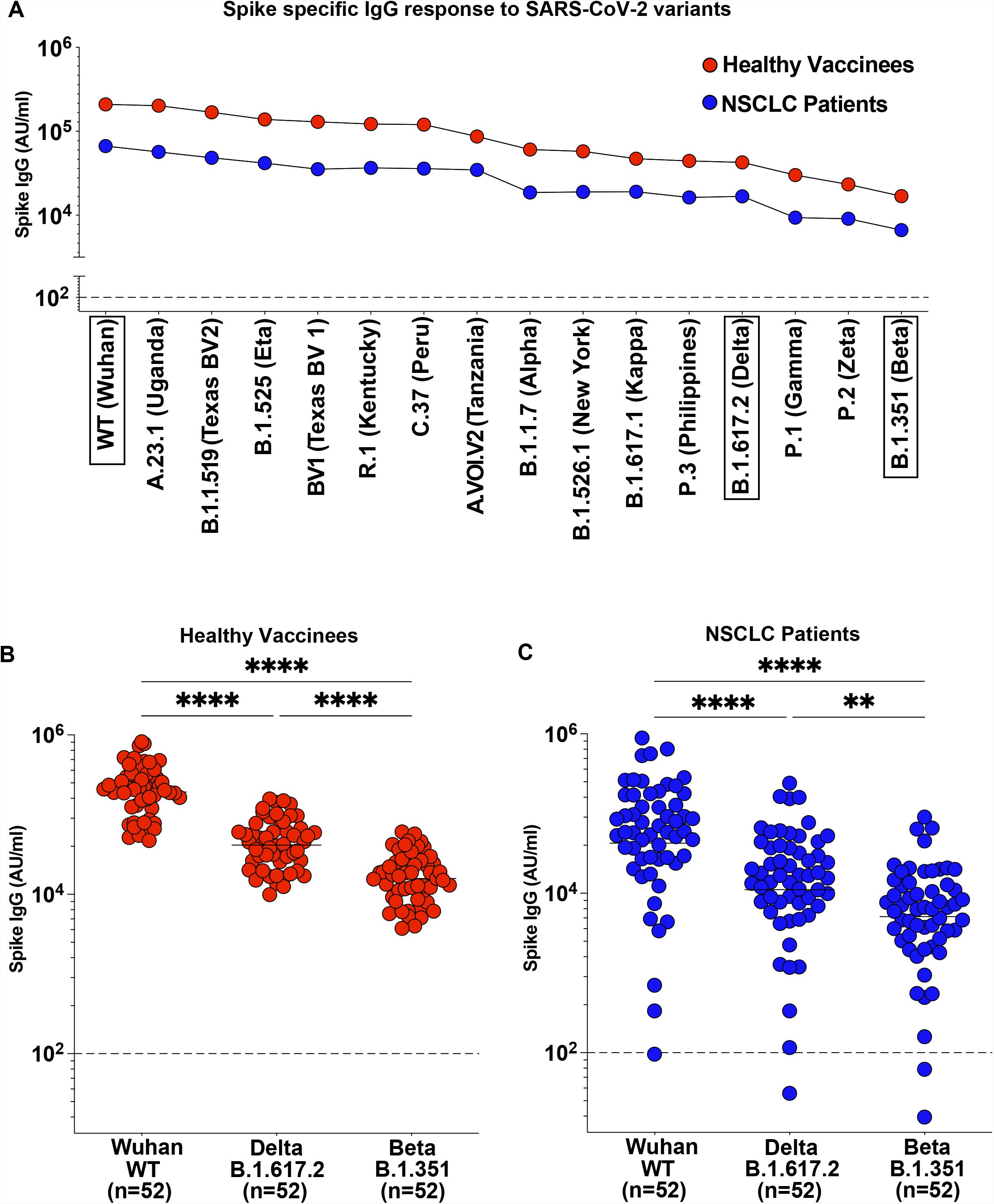
Spike specific IgG response to SARS-CoV-2 variants in NSCLC patients. Figure 5A. Median IgG titers in plasma from NSCLC patients (n=54) in closed blue circles against different variant spike antigens compared to the median IgG titers in healthy (n=52) vaccinees in closed red circles. Figure 4B. Spike-specific IgG titers in healthy vaccinees to WT, Delta and Beta variants in closed red circles. Figure 4C. Spike-specific IgG titers in NSCLC patients to WT, Delta and Beta variants in closed blue circles. Statistical differences were measured using one-way anova. Graph shows the mean and s.e.m. ****p≤0.0001.

### Live-virus neutralizing antibody response against SARS-CoV-2 Delta Beta and Omicron variants in vaccinated NSCLC patients

Next, we performed a live virus neutralization assay to determine if SARS-CoV-2 mRNA vaccination in NSCLC patients induced neutralizing antibodies against the variants of concern including the recently emerged B.1.1.529 (Omicron) variant. NSCLC patient samples with the highest neutralizing antibody titer against the WT strain (in Fig 3A) were used for the VOC neutralization assay. As shown in Fig 6, plasma samples from NSCLC patients had 6.47-fold lower neutralizing capacity against the Delta variant compared to the WT strain (P=0.0001). The neutralizing antibody titer to the Beta variant was 14-fold lower compared to the WT strain (P=<0.0001). More importantly, we observed a significantly lower neutralizing antibody titer against the Omicron variant (79.54-fold) compared to the WT neutralizing antibody titer (P=<0.0001) in NSCLC patients (Fig 6). These data suggests that after a two dose SARS-CoV-2 mRNA vaccination, there is considerable neutralizing activity against the WT virus. However, the capacity of this antibody response to neutralize the Omicron variant is severely limited.

**Figure 6.**
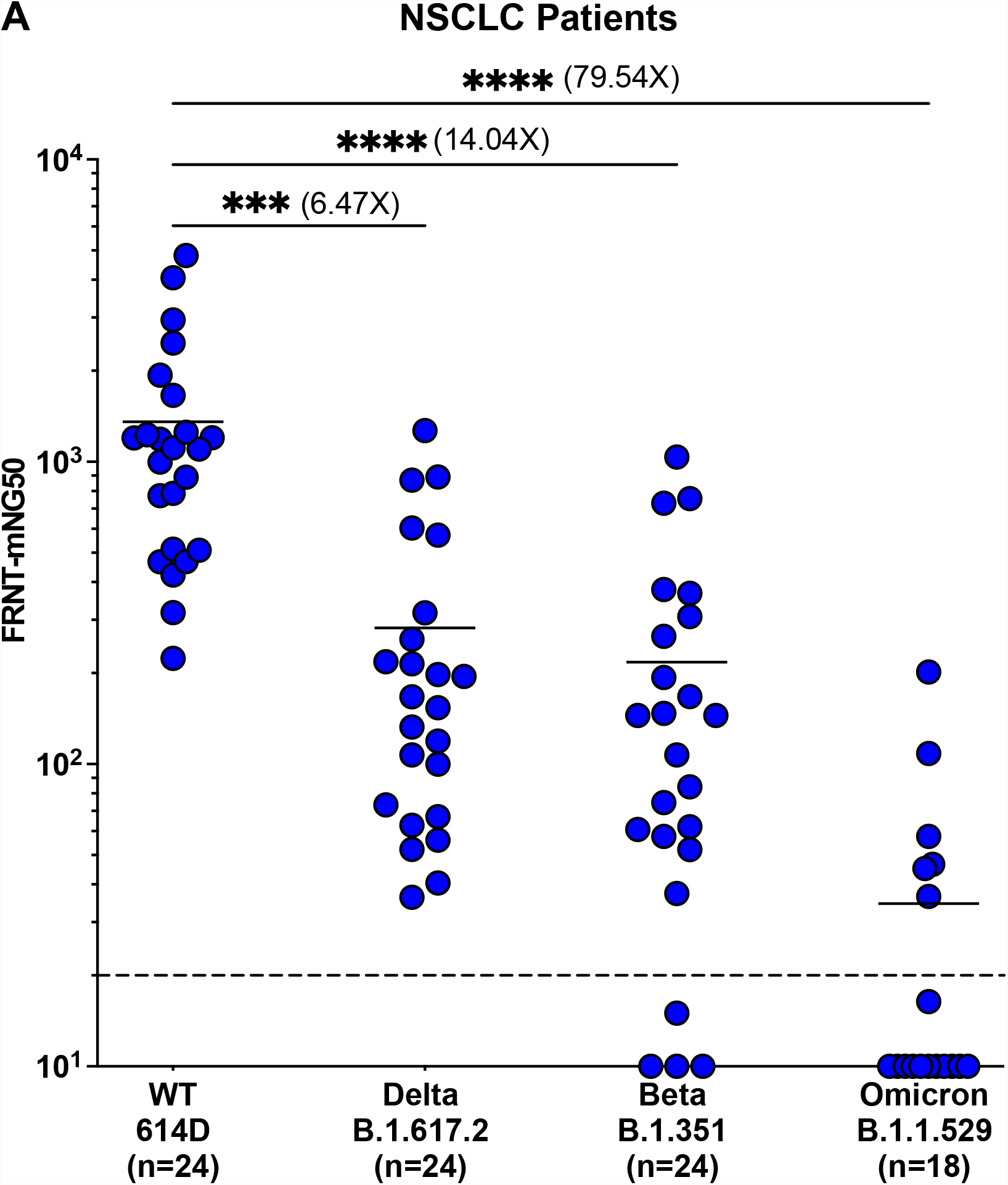
Neutralizing antibody response to Delta, Beta and Omicron SARS-CoV-2 variants in NSCLC patients. Live virus neutralization of WT (614D), B.1.617.2 (Delta), B.1.351 (Beta) and B.1.1.529 (Omicron) virus by sera from NSCLC patients. Statistical differences were measured using one-way anova. Graph shows the mean and s.e.m. ****p≤0.0001.

## DISCUSSION

Immunosuppression in cancer patients increases susceptibility to infections and reduces response to vaccination ^24,25^. Recent studies show that a subset of cancer patients with solid and hematologic malignancies fail to respond to SARS-CoV-2 mRNA vaccination^26-28^. Numerous SARS-CoV-2 variants of concern are currently evolving which evade vaccine-induced antibody response even in vaccinated healthy individuals ^29^. This leaves cancer patients with immune dysfunction at a higher risk to contract severe SARS-CoV-2 disease.

Here, we studied the binding and neutralizing antibody response to SARS-CoV-2 mRNA vaccination in NSLC patients compared to healthy individuals. Our study focusses especially on the antibody response in NSCLC patients to SARS-CoV-2 variants of concern. We report that though most NSCLC patients generate a vaccine specific antibody response, their binding and neutralizing antibody titers are significantly lower compared to healthy controls. Severe immune dysregulation could be a factor for poor vaccine-induced antibody responses in these patients. Vaccine-induced anti-spike and RBD antibody titers in NSCLC patients were significantly reduced after six months of vaccination. Though the SARS-CoV-2-specifc antibody response persists in vaccinated healthy individuals, the antibody titers decrease significantly over time^30-34^. A booster dose is advised to counter the declining immunity and it has proven to be efficient in boosting binding and neutralizing antibody titers^35^. Demographic factors play a major role in influencing vaccine responses^36^. The humoral response to SARS-CoV-2 vaccination in the elderly is known to be significantly lower compared to young adults ^12,37,38^. NSCLC patients above 70 years of age had significantly lower anti-spike antibody titers compared to patients less than 60 years. Further studies are required to determine if the higher median age of NSCLC patient cohort compared to the healthy vaccinee cohort influenced the lower antibody response in these patients. In NSCLC patients, PD-1 targeted therapies improve the durable response rate and increases long-term survival of the patient ^39^. Combination of chemotherapy and PD-1 targeted therapy results in a higher response rate and prolonged survival^40^. Cancer therapies are known to impair immune responses to infection and vaccination ^24,25^. However, we did not see a significant difference in the antibody response to SARS-CoV-2 vaccination in patients receiving either PD-1 monotherapy or a combination of chemotherapy and PD-1 targeted therapy compared to patients receiving no cancer therapy at the time of vaccination. Further investigation with a larger cohort has to be done to validate this finding.

Several SARS-CoV-2 variants have emerged during the course of the pandemic and some of these variants have acquired mutations that result in enhanced virus transmission and pathogenicity and are not neutralized efficiently by vaccine-induced antibodies^15^. Of note, the delta variant identified in December 2020, emerged as the predominant SARS-CoV-2 strain in the U.S and several parts of the world by August 2021^41^. Our data show that the vaccinated NSCLC patients have significantly lower binding and neutralizing antibody response to the Delta and Beta SARS-CoV-2 variants. The recently emerged Omicron variant, first identified in South Africa and Botswana, is efficient in evading vaccine-induced neutralization in healthy individuals^17,20^. This could potentially be a major concern for cancer patients who have significantly lower neutralizing antibody titers against the wild-type SARS-CoV-2 strain compared to healthy adults. Our data show dramatically reduced neutralization of the Omicron variant by sera from NSCLC patients who received two doses of the mRNA vaccination, suggesting that cancer patients might be more vulnerable to infection with the Omicron variant compared to healthy vaccinated individuals.

## Data Availability

All data produced in the present study are available upon reasonable request to the authors

## Figure Legend

**Supplementary Figure 1**.

Supplementary figure 1 A-C. Correlation between Spike-specific IgG (A), IgM (B) and IgA (C) titers and RBD-specific titers in the plasma of NSCLC patients. Supplementary figure 1 D-F. Correlation of Spike-specific IgG (D), IgM (E) and IgA (F) titers and NTD-specific titers in the plasma of NSCLC patients.

