## Supplementary figures and images for "Antibody response to SARS-CoV-2 mRNA vaccine in lung cancer patients: Reactivity to vaccine antigen and variants of concern"

### Supplemental figure 1

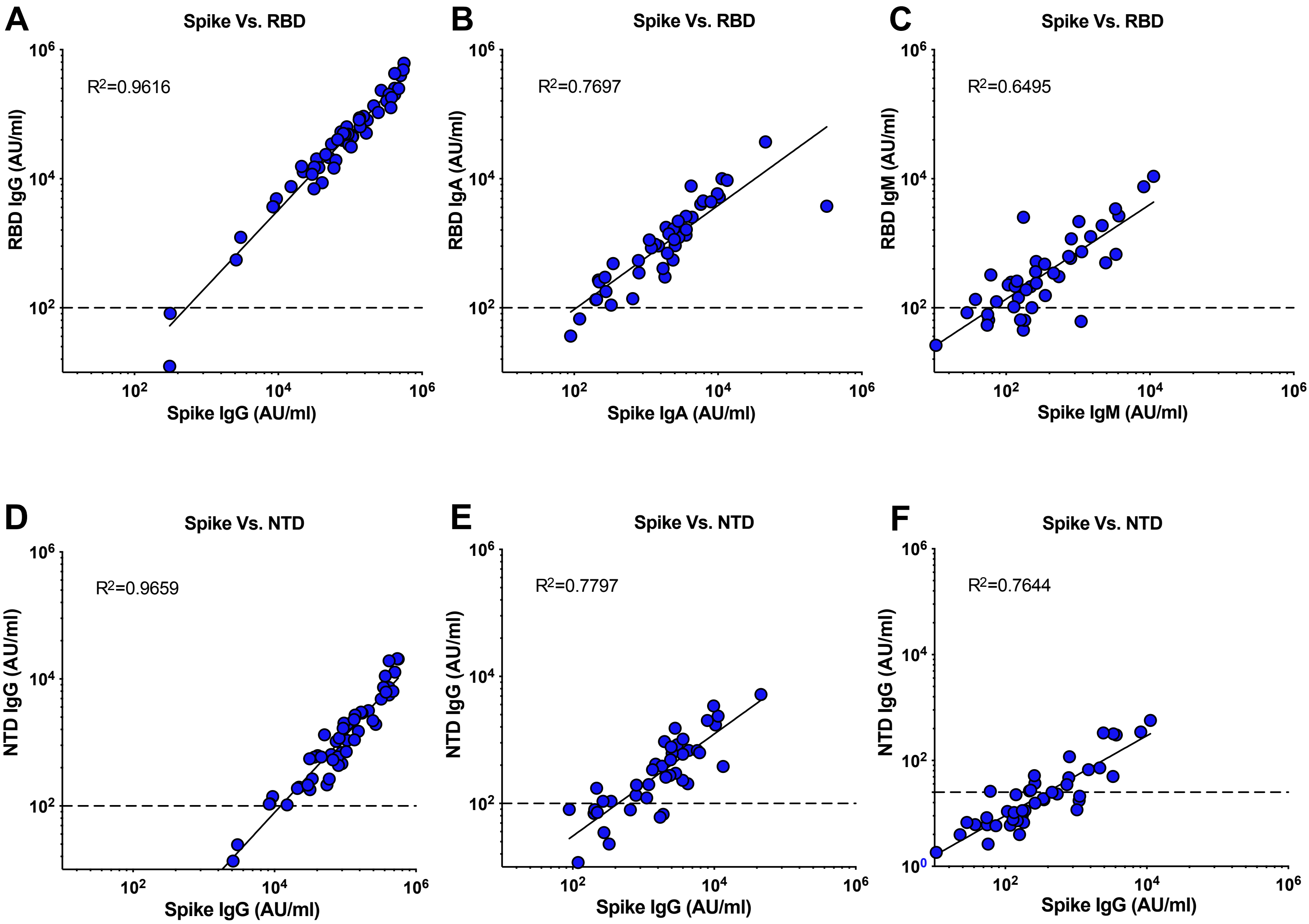


Supplementary Figure 1
